# Massive social protests amid the pandemic in selected Colombian cities: Did they increase COVID-19 cases?

**DOI:** 10.1101/2021.06.16.21258989

**Authors:** José Moreno-Montoya, Laura A. Rodríguez-Villamizar, Alvaro J. Idrovo

## Abstract

**Background:** Since April 28, 2021, in Colombia there are social protests with numerous demonstrations in various cities. The aim of this study was to assess the effect of social protests on the number and trend of the confirmed COVID-19 cases in some selected Colombian cities where social protests had more intensity.

**Methods:** We performed and interrupted time-series analysis (ITSA) and Autoregressive Integrated Moving Average (ARIMA) models, based on the confirmed COVID-19 cases in Colombia, between March 1 and May 15, 2021, for Bogotá, Cali, Barranquilla, Medellín, and Bucaramanga. The ITSA models estimated the effect of social demonstrations on the number and trend of cases for each city by using Newey-West standard errors. ARIMA models assessed the overall pattern of the series and effect of the intervention. We considered May 2, 2021, as the intervention date for the analysis, five days after social demonstrations started in the country.

**Findings:** During the study period the number of cases by city was 1,014,815 for Bogotá, 192,320 for Cali, 175,269 for Barranquilla, 311,904 for Medellín, and 62,512 for Bucaramanga. Heterogeneous results were found among cities. Only for the cities of Cali and Barranquilla statistically significant changes in trend of the number of cases were obtained after the intervention: positive in the first city, negative in the second one. None ARIMA models show evidence of abrupt changes in the trend of the series for any city and intervention effect was only significant for Bucaramanga.

**Interpretation:** Social protests had a heterogeneous effect on the number and trend of COVID-19 cases. Different effects might be related to the epidemiologic moment of the pandemic and the characteristics of the social protests. Assessing the effect of social protests within a pandemic is complex and there are several methodological limitations. Further analyses are required with longer time-series data.

## Introduction

Physical distancing, use of masks, frequent hand washing, and good ventilation are some of the most effective non-pharmacological measures to prevent the transmission of SARS-CoV-2 which causes Coronavirus Disease (COVID-19).^1^ When people meet, it is difficult to maintain physical distancing, so the recommendations have been to avoid religious services, sports and cultural activities in closed settings, and to control entry into shopping centers, supermarkets and other indoor environments. However, sanitary recommendations are hard to meet when social disorder arises, and people oppose or dispute the government’s decisions.^2^

During the pandemic a lot of social protests have been observed around the world. According to the “COVID-19 Disorder Tracker”, during the pandemic most social protests have occurred in India, Israel and Mexico,^3^ but the best known were that arose in the United States after the murder of George Floyd by a police officer.^4^ In all these cases there was concern about the possible effect of social protests on the transmission of SARS-CoV-2. The few studies that have analyzed whether social protests increase the transmission of SARS-CoV-2 show contradictory results,^5-7^ although the evidence tends to point out that there are no significant increases.

The region of the Americas has been the most affected region during the COVID-19 pandemic with more than 69,8 million cases and 1,8 million deaths up to and including June 14, 2021.^8^ In Colombia, 3,777,600 confirmed cases and 96,366 deaths have been reported up to the same date.^9^ The country is transitioning the third wave of the epidemic which has registered the highest counts of daily reported cases and deaths. In the middle of this third wave, a large social movement with social protests arouse across the country by the end of April. This social national movement is known as “Paro Nacional” and the first social protests took place across different cities on April 28, 2021, and, although intensity has decreased, it has been active till until when this report is written (June 15, 2021).

Some social scientists have indicated that the main causes of the social protests are social discontent, increase in poverty and unemployment, a tax reform proposal (withdrawn by the government as a result of social protest), a health reform, criticism of management of pandemic (mainly due to delay and slow vaccination), and excessive police force against protesters. Other problems noted are government non-compliance with the peace process and the increase in violence in some regions.^10^ The social discontent turned out to be more important than the perception risk of infection with SARS-CoV-2 among an important proportion of Colombians.^11^

These social protests started when several cities had the highest occurrences of new cases of COVID-19 and intensive care units were in short supply. The potential effect of social protests, if present, might represent an increase in the number of new cases; however, to our knowledge no epidemiological studies has been conducted to assess it. In this context, this study aimed to assess the effect of social protests on the number and trend of the confirmed COVID-19 cases during the third wave of the epidemic in five selected Colombian cities.

## Material and methods

We conducted a time-series ecological study using the daily counts of COVID-19 cases before and after the social protests that started on April 28, 2021, in selected Colombian cities: Bogotá, Barranquilla, Bucaramanga, Cali, and Medellín. These cities were selected for their high risk of SARS-CoV-2 transmission among citizens, consequence of their high population density, and because in these cities the social protest had more intensity. In this sense, if social protests had effect on the incidence of COVID-19 cases is expected to be more evident in these cities. This study follows a quasi-experimental approach with a single group (uncontrolled) before- and after-design.

The daily number of confirmed cases of SARS-CoV-2 infection were obtained for Colombia from the official COVID-19 website of the National Institute of Health (https://www.ins.gov.co/Noticias/Paginas/coronavirus-casos.aspx) on June 6, 2021, including all reported cases up to June 5, 2021. For defining the final point for the analysis, we considered the delay in the report of the cases. The delay between the date of symptoms onset (DSO) and the report of cases to the public health surveillance system during 2021 has a median of 10 days (interquartile range 6-16 days) and 95% percentile at 22 days. Then, reliable reported data (at least 95% of reported cases) for the DSO is available in the dataset up to 21 days before the cut-off date (May 15, 2021). Hence, we used the daily count of confirmed cases by DSO, between March 1 and May 15, 2021, as the dataset for analysis.

Cases are confirmed in the National Surveillance System for Public Health (SIVIGILA, for its name in Spanish) by using Real-Time Protein Chain Reaction (RT-PCR) or antigen tests. We used the DSO as the incidence date for COVID-19 cases due to the date of contagion is unknown. For asymptomatic confirmed cases we assigned the date of diagnosis as the DSO. The epidemic curve for Colombia using the DSO shows that in the current third wave of the epidemic, the daily number of cases start to rise early in March (Supplementary Figure 1). Thus, we used March 1, 2021, as the initial point for our analysis as we aimed to assess the potential effects of social protests on the current epidemic wave.

### Statistical methods

COVID-19 daily data was considered as a time-series. In consequence, the data are considered to follow a relative deterministic pattern with some degree of autocorrelation. We used two analytical approaches: interrupted time-series analysis (ITSA) by means of Newey-West regressions of robust autoregressive errors and Autoregressive Integrated Moving Average (ARIMA) models.

ITSA assessed the potential effects of social protest as step “intervention”. For this purpose, we divided the time-series into two moments, before and after the social protest in Colombia under the hypothesis that social protests might affect the structure of time variation in the number of cases. The ITSA is a special case of time-series design that assumes that the intervention occurs at a specific point in time, the outcome is expected to change immediately and abruptly as a result of the intervention and the pre-intervention period is assumed as the counterfactual.^12^ For defining the intervention point for the analysis we used the COVID-19 incubation period which is estimated to be around 2-14 days with a mean of 5-7 days.^13^

The potential effects of social protests, if present, might be reflected in DSO on average 5-7 days after they started. The start date of social protests was on April 28, 2021, when the first national social protests were registered across the country. Then, potential effects of social protests on DSO, if present, should be reflected in incident cases with DSO on average after five days on May 2, 2021. Therefore, we used May 2, 2021, as the intervention date for the analysis. We used a standard single-group ITSA using ordinary least square regression model with Newey-West standard errors and autocorrelation at lag 1. The model calculates the coefficient for the slope prior to intervention (β1), the coefficient of change in level in the time immediately after the intervention compared to counterfactual (β2), and the coefficient for the difference between pre- and post-intervention slopes (β3). The coefficient for the post-intervention trend (β4) is estimated using a linear combination of estimators.^14^ We used the Cumby and Huizinga general test for assessing the autocorrelation in the time series. We conducted an alternative analysis using a shortened time-series preintervention period (April 12 – May 15, 2021) to assess the effect of protests considering the two weeks period before the intervention to rule out the long-trend effect of the third wave of the pandemic.

ARIMA models’ approach was also used to assess the overall time pattern of the series. It differs from ITSA methodology because it does not consider a specific trend in the historical data of the sequence to be analyzed. Instead, it uses an interactive approach to identify a possible model from a general model class. The chosen model is then tested against historical data to see if the sequence is correctly represented.

In absence of an intervention, the total of cases of COVID-19 could be explained by the ARIMA model and based exclusively in the historical information from the number of cases. In such case, residuals of the model are not expected to show a particular pattern of variation and thus, having a white noise behavior. In cases of interventions, the series could be nonstationary and nonlinear, going from one status to another in a complex manner. An intervention variable was included in each model considering the same date than the ITSA analysis. Auto-correlogram (ACF) and partial auto-correlogram (PACF) functions were analyzed for each city for the model choice and Ljung–Box portmanteau (Q) test for white noise for examining the residuals^15,16^. The candidate models were compared using BIC selection criteria^16^. We conducted all analyses separately for each city. We used the Stata version 16 and the R software.

## Results

There was a total of 3,547,017 confirmed COVID-19 cases in Colombia up to and including June 5^th^. The total number of confirmed cases by city were 1,014,815 for Bogotá, 192,320 for Cali, 175,269 for Barranquilla, 311,904 for Medellín, and 62,512 for Bucaramanga. Figure 1-A, B, C, D, E presents the description of the time-series of confirmed cases by DSO for each city between March 1 and May 15, 2021.

**Figure 1.**
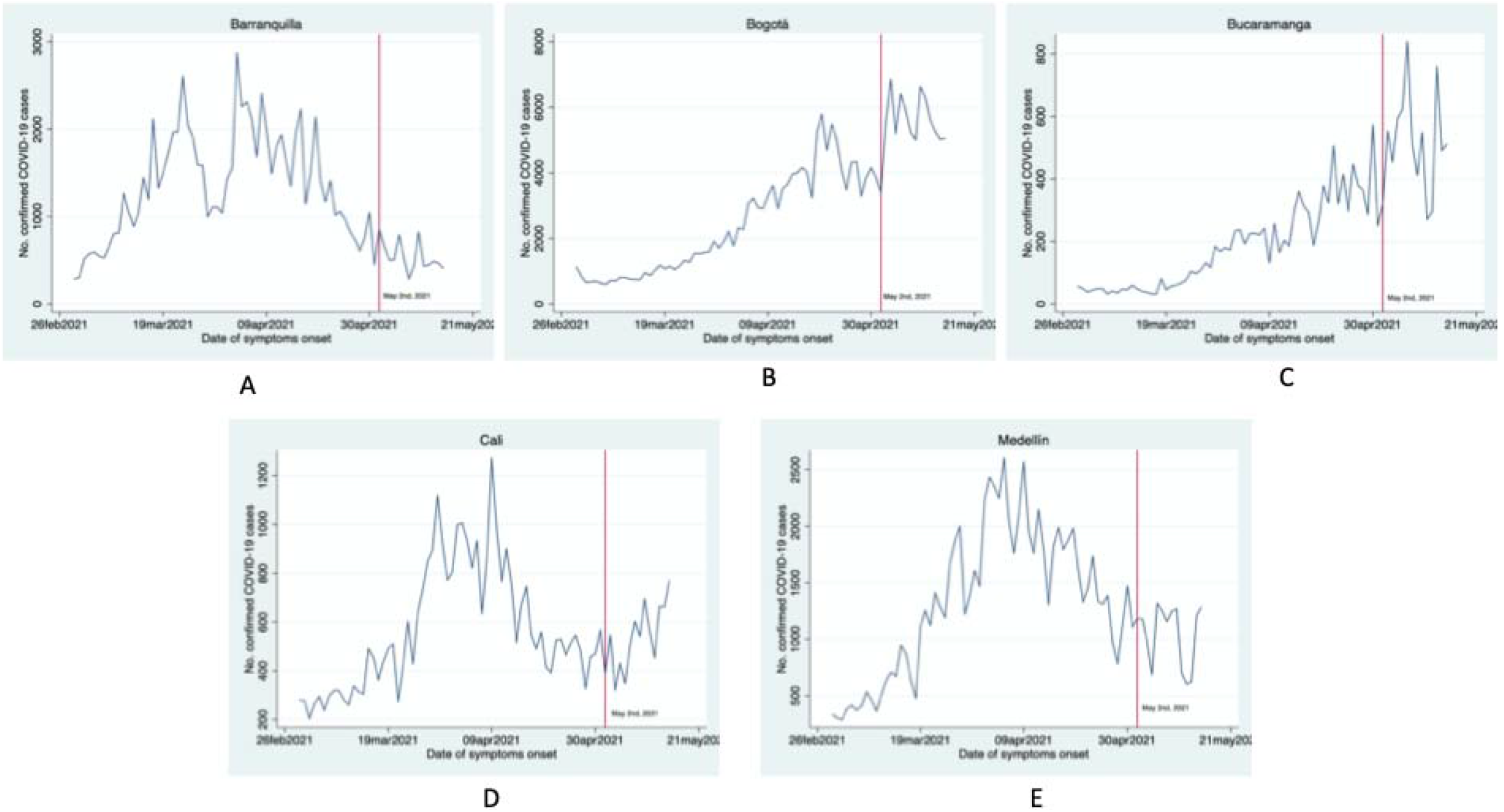
Time-series of COVID-19 confirmed cases in selected cities during the period studied (March 1 to May 15, 2021).* A) Barranquilla; B) Bogotá; C) Bucaramanga; D) Cali; E) Medellín The vertical red line corresponds to May 2, 2021, the selected “intervention” date.

### ITSA Analysis

Table 1 and Figure 2 present the results of ITSA models for all cities during March 1 and May 15, 2021. For Bogotá and Bucaramanga, in the first day after the intervention date (April 2, 2021), there appeared to be an increase in the number of cases by DSO (β1) while in Barranquilla, Cali, and Medellín the number of cases decreased; however, only coefficients for these three cities were statistically significant. The daily trend of cases compared to the pre-intervention period (β2) and the trend in the post-intervention period (β3) was positive for Cali and negative for Barranquilla, both with statistical significance. Table 1 shows the results of the alternative analysis using the time series between April 12 and May 15, 2021. For all cities except for Cali, in the first day after the intervention date (April 2, 2021), there appeared to be an increase in the number of cases by DSO (β1); however, all coefficients were not statistically significant. For Barranquilla and Cali, the daily trend of cases compared to the pre-intervention period (β2) was positive and statistically significant. The trend in the post-intervention period (β3) was positive for Cali and negative for Barranquilla, both with statistical significance.

**Table 1.** Results of interrupted time-series analysis for five Colombian cities.

| City | March 1 to May 15, 2021 |  |  | April 12 to May 15, 2021 |  |  |
| --- | --- | --- | --- | --- | --- | --- |
| | $\beta_1$ (95% CI) | $\beta_2$ (95% CI) | $\beta_3$ (95% CI) | $\beta_1$ (95% CI) | $\beta_2$ (95% CI) | $\beta_3$ (95% CI) |
| Barranquilla | -903.36<br>(-1384 to -422,11) | -27.27<br>(-44.75 to -9.80) | -20.30<br>(-34.22 to -6.38) | 152.03<br>(-114.04 to 418.10) | 50.37<br>(24.33 to 76.41) | -20.30<br>(-35.08 to -5.52) |
| Bogotá | 639.86<br>(-663 to 1942.84) | -64.85<br>(-212.66 to 82.94) | 12.17<br>(-133.97 to 158.32) | 1218.98<br>(-166.85 to 2604.82) | 9.75<br>(-157 to 176.53) | 12.17<br>(-142.96 to 167.31) |
| Bucaramanga | 126.37<br>(-14.35 to 267.09) | -7.24<br>(-24.53 to 10.05) | -0.55<br>(-17.68 to 16.56) | 96.98<br>(-45.53 to 239.49) | -8.90<br>(-29.62 to 11.80) | -0.56<br>(-18.73 to 17.61) |
| Cali | -360.92<br>(-545.70 to -176.16) | 19.04<br>(8.08 – 29.99) | 24.37<br>(14.10 to 34.64) | -16.35<br>(-155.41 to 122.70) | 38.77<br>(23.21 to 54.34) | 24.37<br>(13.46 to 35.28) |
| Medellín | -884.71<br>(-1360 to -408.55) | -32.68<br>(-71.97 to 6.60) | -11.13<br>(-49.23 to 26.96) | 103.10<br>(-251.55 to 457.73) | 36.37<br>(-10.77 to 83.51) | -11.13<br>(-51.58 to 29.31) |

$\beta_1$ = Coefficient for the effect in level of cases in the day after intervention
$\beta_2$ = Coefficient for daily trend of cases in post-intervention compared to the pre-intervention period
$\beta_3$ = Coefficient for the trend in the post-intervention period

**Figure 2.**
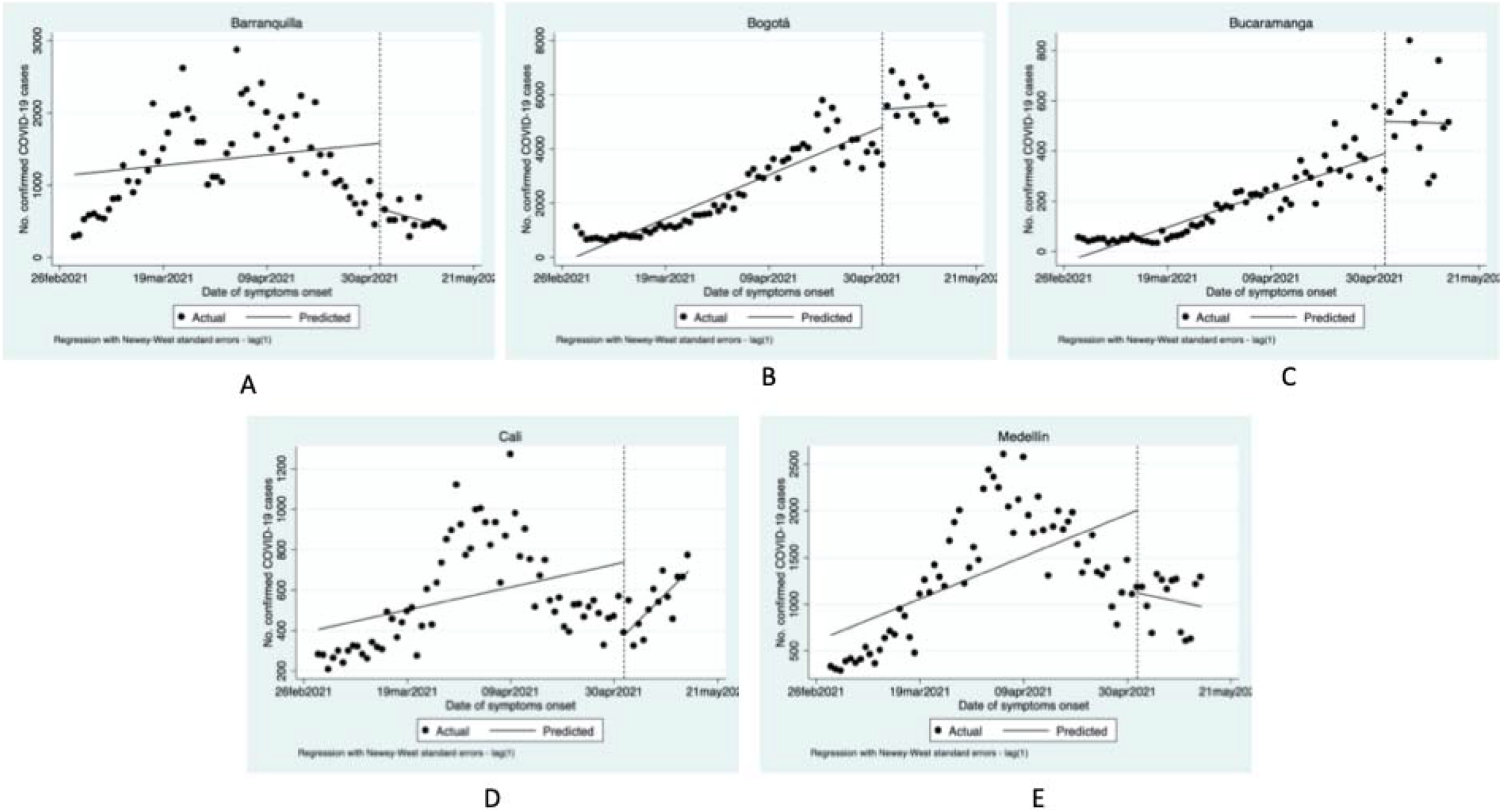
Interrupted time-series analysis plots for five capital cities with series data between March 1 and May 15, 2021. A) Barranquilla; B) Bogotá; C) Bucaramanga; D) Cali; E) Medellín

### ARIMA models

The analyzed dates ranged from March 1 to May 15, 2021. In the identification of the model, the ACF and PACF were inspected in COVID-19 confirmed cases (Figure 3). A linear trend pattern was corrected by first degree order of differentiation for all the cities analyzed excepting for Medellin who had a more marked growing trend with an integration pattern of degree 2.

**Figure 3.**
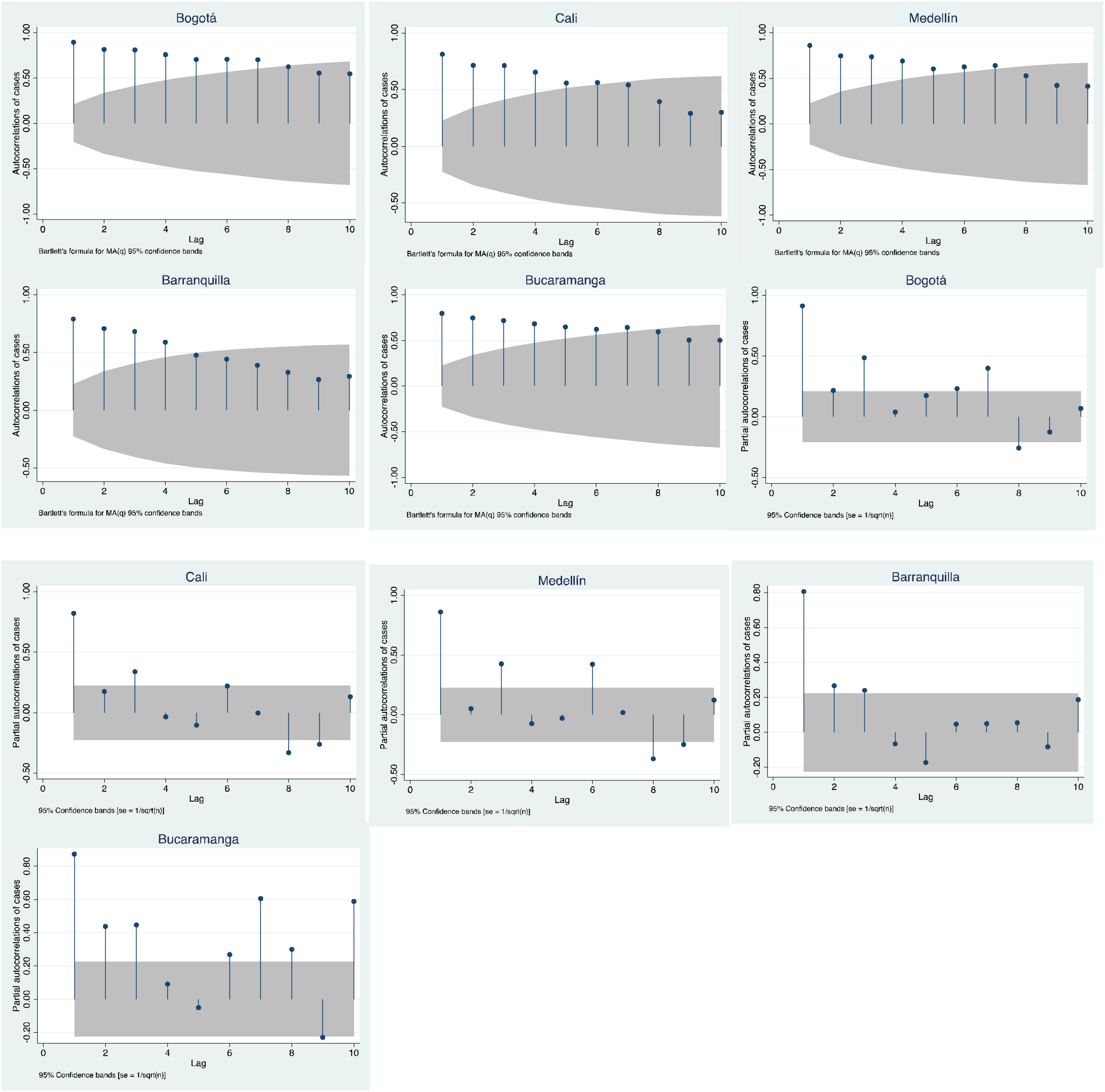
Auto-correlations and partial auto-correlations for five selected cities with series data between March 1 and May 15, 2021.

A consistent autoregressive behavior was verified in all analyzed cities as well as a trend pattern. The number of lags considered varied from 7 days in Bogotá to 2 days in Cali. Due differentiation, all estimated coefficients were negative in the auto-regressive part of the models, so it is expectable to see an increasing trend in the series (Figure 1). Except by Medellín, no moving-average effects were appreciated in the series, i.e., just autoregressive models were adjusted. After ARIMA models, no residuals rejected the white noise assumption, what can be interpreted as the inexistence of abrupt changes in the time series pattern (Table 2). Intervention effects were discarded for four of the analyzed cities except for Bucaramanga where a positive significant coefficient was estimated (p=0.008, see table 2).

**Table 2.**
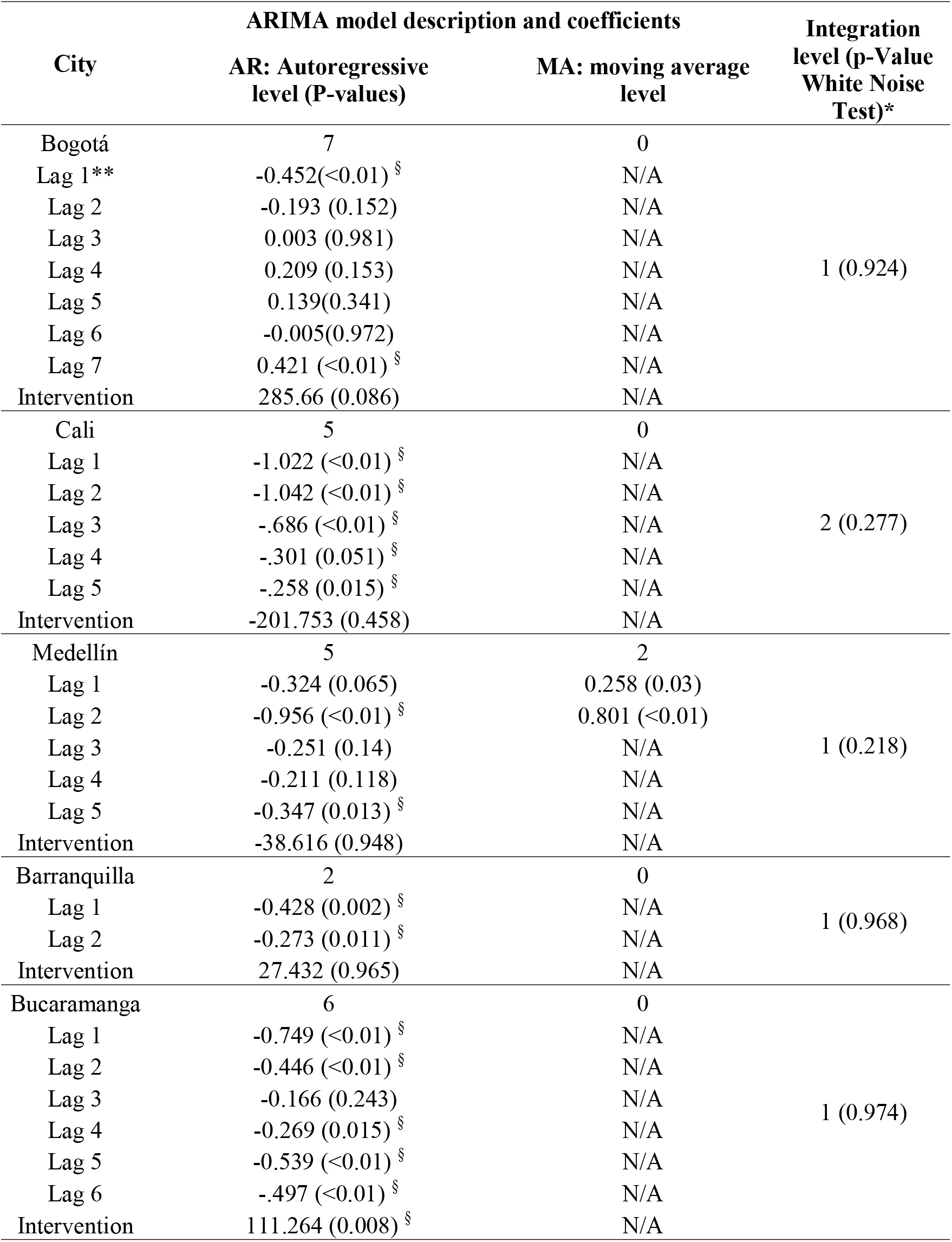
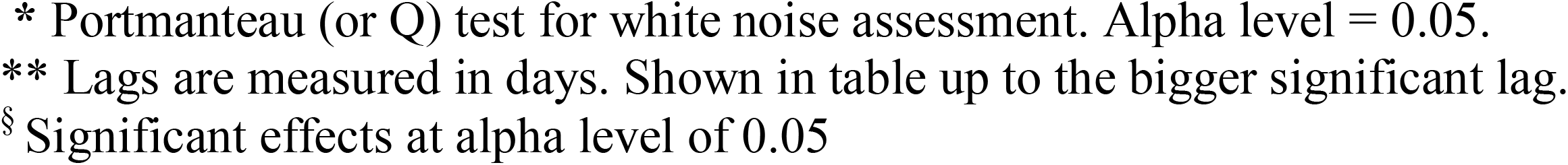
Results of ARIMA models for five Colombian cities. March 1 to May 15, 2021.

| City | ARIMA model description and coefficients |  | Integration level (p-Value White Noise Test)* |
| --- | --- | --- | --- |
|  | AR: Autoregressive level (P-values) | MA: moving average level |  |
| Bogotá | 7 | 0 | 1 (0.924) |
| Lag 1** | -0.452(<0.01) § | N/A |  |
| Lag 2 | -0.193 (0.152) | N/A |  |
| Lag 3 | 0.003 (0.981) | N/A |  |
| Lag 4 | 0.209 (0.153) | N/A |  |
| Lag 5 | 0.139(0.341) | N/A |  |
| Lag 6 | -0.005(0.972) | N/A |  |
| Lag 7 | 0.421 (<0.01) § | N/A |  |
| Intervention | 285.66 (0.086) | N/A |  |
| Cali | 5 | 0 | 2 (0.277) |
| Lag 1 | -1.022 (<0.01) § | N/A |  |
| Lag 2 | -1.042 (<0.01) § | N/A |  |
| Lag 3 | -.686 (<0.01) § | N/A |  |
| Lag 4 | -.301 (0.051) § | N/A |  |
| Lag 5 | -.258 (0.015) § | N/A |  |
| Intervention | -201.753 (0.458) | N/A |  |
| Medellín | 5 | 2 | 1 (0.218) |
| Lag 1 | -0.324 (0.065) | 0.258 (0.03) |  |
| Lag 2 | -0.956 (<0.01) § | 0.801 (<0.01) |  |
| Lag 3 | -0.251 (0.14) | N/A |  |
| Lag 4 | -0.211 (0.118) | N/A |  |
| Lag 5 | -0.347 (0.013) § | N/A |  |
| Intervention | -38.616 (0.948) | N/A |  |
| Barranquilla | 2 | 0 | 1 (0.968) |
| Lag 1 | -0.428 (0.002) § | N/A |  |
| Lag 2 | -0.273 (0.011) § | N/A |  |
| Intervention | 27.432 (0.965) | N/A |  |
| Bucaramanga | 6 | 0 | 1 (0.974) |
| Lag 1 | -0.749 (<0.01) § | N/A |  |
| Lag 2 | -0.446 (<0.01) § | N/A |  |
| Lag 3 | -0.166 (0.243) | N/A |  |
| Lag 4 | -0.269 (0.015) § | N/A |  |
| Lag 5 | -0.539 (<0.01) § | N/A |  |
| Lag 6 | -.497 (<0.01) § | N/A |  |
| Intervention | 111.264 (0.008) § | N/A |  |

- \* Portmanteau (or Q) test for white noise assessment. Alpha level = 0.05. - \*\* Lags are measured in days. Shown in table up to the bigger significant lag. - § Significant effects at alpha level of 0.05

## Discussion

Results of these analyses suggest that there were different effects in the cities included in the study. Cali and Bucaramanga were the cities where our results suggest an effect of social protests on the incidence of COVID-19 cases, from the ITSA and ARIMA model, respectively, but not from both at the same time. In the other cities there were no “a peak within the peak”,^17^ as it had been informed in Colombia in the midst of the social protests. Given the heterogeneity of the results, it is needed to analyze each city in isolation to have a better interpretation of the findings. In Cali the third wave had already passed when protests began, and it seems the number of incident cases of SARS-CoV-2 infection started to increase again. In Barranquilla and Medellín the highest number of cases reached during the third wave had already been surpassed when the social protests began, and the trend in the number of cases was stable after protests. In Bogotá and Bucaramanga, the data suggest that before the social protests there was already an increase in the number of cases. For this reason, the risk of social protests-related adverse effects was higher in these cities if the hospitals had already a high occupancy of patients, as occurred in Bucaramanga.

In all cities, social protests were outdoor activities with different occurrence of use of masks and compliance with biosafety standards. Unfortunately, there are no objective data to clarify this important issue. However, in Cali there were large marches even with strong confrontations between protesters and police, and previous data suggests that the surveillance data is reliable.^18^ Perhaps these two facts made it possible to identify the effect of social protests on the trend of incident cases. The data on the decline of the wave observed in Barranquilla and Medellín perhaps prevented the identification of the effects of social protest. It is even possible that public health surveillance slows down its rate of finding cases.

These findings can be compared with the results obtained in studies on social protests after the George Floyd death in USA.^5-7^ Three studies available shown contradictory results, although the evidence tends to point out that there are no important increases in the occurrence de COVID-19 cases. The studies include different populations, which can be associated with no-consistent results. However, this fact supports the findings of our study where we also found different results in cities. Although crowds are still risky for the transmission of SARS-CoV-2 there is interesting evidence that challenges this general premise, in specific contexts. A very striking recent study is a clinical trial that assessed the effectiveness of facial masks and adequate air ventilation at a music event in Spain. In this case, there was no evidence of an increase in the number of COVID-19 cases, despite the fact that mass-gathering indoor events are considered high-risk contexts.^19^

The results of this study should be interpreted with caution, considering the limitations inherent in the data and statistical methods used. First, temporal changes in the number of confirmed cases of SARS-CoV-2 infection should not be interpreted as changes in viral transmission. Data available in Colombia do not allow this inference to be made since public health surveillance has not been adequate to identify all the contacts of each new case that is reported. It is consequence of the way in which public health surveillance is carried out, which privilege symptomatic cases and has limitations in contact tracing, as described by Vecino-Ortiz et al.^20^ Thus, confirmed cases might not be representative of transmission due to underreporting. Second, it is not clear whether there was non-differential report during the study time. The different moments of the pandemic in each city make it difficult to identify whether the case identification strategies can be comparable. Available data do not allow a rigorous evaluation of this issue. Third, data used in the analysis only included a two-weeks period after the incubation period and later effects should be evaluated later in time. However, we recognize that it is difficult to test later effects given that there were events, such as the celebration of Mother’s Day (first Sunday of the month; in this year May 9, 2021), that made a rigorous evaluation more difficult. Fourth, most participants in social protests are young people, and violent acts do not usually occur in social protests where many older individuals participate. Therefore, if young people are infected during the protests, they may be asymptomatic and do not seek diagnosis. This underreporting may have made it difficult to identify the association between social protests and the increase in cases. Finally, our assessment does not correspond to a long-time analysis. Structural patterns of the series could be overlooked.

Beyond the results of this study, it is possible that social protests are associated with changes in mortality. A possible direct mechanism is that social protests increase the transmission of SARS-CoV-2, resulting in the occurrence of complicated clinical cases and deaths among those infected. The indirect mechanism is that social protests are associated with injuries that require urgent medical attention, which generates greater use of health services that could be used to treat COVID-19 patients. The possible consequence is that there is no possibility of care in intensive care units, no availability of respirators, which leads to death. However, these possible effects are beyond the analysis presented here.

In conclusion, this study suggests there is heterogeneity in the association between social protests and an increase in the number of incident cases of SARS-CoV-2 across selected cities in Colombia. These divergent results that might be related to the epidemiologic time of the epidemic in each city. Given the inherent risk of social protests, the best scenario to manage the pandemic is that they do not exist. It is urgent that protesters and the government officials negotiate and reach minimum agreements that reduce the risk of COVID-19 transmission and improve the conditions that led to social discontent. These findings can contribute to reduce fake news and to support evidence-based decision making.

## Data Availability

Data used in this study are public. They are available at https://www.ins.gov.co/Noticias/Paginas/coronavirus-casos.aspx

## Notes

### Competing Interest Statement

The authors have declared no competing interest.

### Clinical Trial

NA

### Funding Statement

This study was not specific funding

### Author Declarations

This study did not require IRB approval accoridng to the Colombian legislation (anonymous secondary data).

