## Supplementary material for "Massive social protests amid the pandemic in selected Colombian cities: Did they increase COVID-19 cases?": Figure 1 supp

Supplementary Figure 1. Time-series of COVID-19 confirmed cases in Colombia, 2020-2021

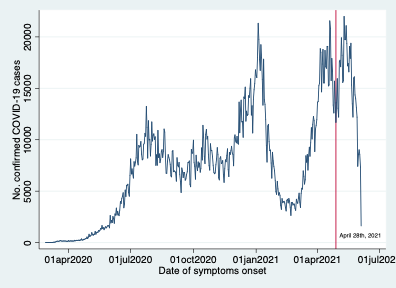

The vertical red line corresponds to April 28, 2021, the first day of social protests in Colombia
